# Long Term Physical, Mental and Social Health Effects of COVID-19 in the Pediatric Population: A Scoping Review

**DOI:** 10.1101/2021.09.17.21263743

**Authors:** Madeline Borel, Luyu Xie, Adrian Mihalcea, Jeffrey Kahn, Sarah E. Messiah

**Affiliations:** University of Texas Health Science Center, School of Public Health, Dallas Campus, Dallas, TX, USA; Center for Pediatric Population Health, UTHealth School of Public Health and Children’s Health System of Texas, Dallas, TX, USA; University of Texas Health Science Center, School of Public Health, Houston Campus, Houston, TX, USA; University of Texas Southwestern Medical Center, Department of Pediatrics, Dallas, TX, USA; Children’s Health System of Texas, Dallas, TX USA

## Abstract

**Background:** The majority of COVID-19 symptom presentations in adults and children appear to run their course within a couple of weeks. However, a subgroup of adults has started to emerge with effects lasting several months or more after initial infection. However, little is known about long term physical, mental and social health effects of COVID-19 in the pediatric population. The purpose of this review was to determine these impacts well into the second year of the pandemic.

**Methods:** A search was conducted using PUBMED, Web of Science, Science Direct, and COCHRANE between 11/1/2019 and 9/1/2021. Search inclusion criteria were as follows: (1) COVID-19 illness and symptoms in children; (2) SARS-COv2 in children; (3) English language; and (4) human studies only.

**Results:** The few studies that have documented long-term physical symptoms in children show that fatigue, difficulty in concentrating (brain fog), sleep disturbances, and sensory problems are the most reported outcomes. Most studies examining the impact of COVID-19 in pediatric populations have focused on initial clinical presentation, and symptoms, which are similar to those in adult populations. Additionally, COVID-19 has had a moderate impact on children and adolescents’ social environment, which may exacerbate current and future physiological, psychological, behavioral, and academic outcomes.

**Conclusions:** There are limited studies reporting long physical symptoms of COVID-19 in the pediatric population. However, pediatric COVID-19 cases are underreported due to low rates of testing and symptomatic infection, which calls for more longitudinal studies. Children who have experienced COVID-19 illness should be monitored for long physiological, psychological, behavioral, and academic outcomes.

## INTRODUCTION

Severe acute respiratory syndrome coronavirus 2 (SARS-CoV-2) causes the novel coronavirus disease 2019 (COVID-19) and as of September 9, 2021, there have been 5.3 million reported cases among children in the United States [1]. Early studies from China [2-9] and Europe [10] have shown that COVID-19 is generally a mild disease in children, including infants. In the US and globally, fewer cases of COVID-19 have been reported in children (age 0-17 years) compared with adults [1][7] but recent data suggests the delta (B.1.617.2) variant is more transmissible among children compared to the alpha (B.1.1.7) variant. Specifically, while children comprise 22% of the US population (novel coronavirus) recent data show that 15.5% of all cases of COVID-19 reported to the CDC were among children [3]. Indeed, the true incidence of SARS-CoV-2 infection in children is not known due to lack of widespread testing and the prioritization of testing for adults and those with severe illness. Also, hospitalization rates in children have remained significantly lower than adult rates suggesting that children may have less severe illness from COVID-19 compared to adults [9]. However, a small proportion of children develop severe disease requiring ICU admission and prolonged ventilation [11], although fatal outcome is rare. Also, reports of a novel Kawasaki disease–like multisystem inflammatory syndrome (MIS-C) necessitate continued surveillance in pediatric patients [12]. This syndrome has also been reported as Pediatric Inflammatory Multisystem Syndrome Temporally Associated with SARS-CoV-2 (PIMS-TS) [13-14].

Common acute symptoms of COVID-19 disease in both adults and children include fever, cough, shortness of breath, chills, muscle pain, headache, sore throat, and loss of taste or smell. The most common physical symptoms reported by adults after a SARS-CoV-2 infection are fatigue/lethargy and shortness of breath, with on average one-third reporting at least one persistent symptom months after recovery [15,16]. Most patients recover within two weeks of initial symptoms. However, a sub-group of adults has been documented to have longer-lasting symptoms, often termed “long” or “long haulers”. One recent study has reported similar long symptoms in children (≤18 years old) previously diagnosed with COVID-19 from the largest hospital in Rome [17]. A questionnaire was delivered by two pediatricians, either online or during outpatient visit, between September 1^st^, 2020 and January 1^st^, 2020 to the children’s caregivers. Symptoms frequently reported up to 120 days after infection included muscle and joint pain, headache, insomnia, respiratory issues, and palpitations [17]. While this study did provide some insight into long COVID-19 symptoms, the findings were limited by a single-center design and relatively small sample size.

The COVID-19 pandemic and its associated mitigation strategies are expected to have significant psychosocial, behavioral, socioeconomic, and health impacts, which are exacerbated in populations that experience health disparities and other vulnerable groups.[18-20] Pediatric populations experiencing health disparities prior to the COVID-19 pandemic are at increased risk of infection and other COVID-19 related consequences (e.g., prolonged school closings, low resources to support online learning, parent job loss, high prevalence of community morbidity and mortality due to COVID-19) [21,23]. Preliminary reports in the U.S. point consistently to disparities by race and ethnicity, with African Americans, Hispanics/Latinos, American Indians/Alaska Natives, and Native Hawaiians/Other Pacific Islanders experiencing a greater COVID-19 burden than non-Hispanic White populations [24]. Reports by geographic locations indicate that cases are substantially greater in economically disadvantaged census tracts [1][25]. These long-term effects, even if only mild in severity, can have a detrimental impact on a person’s overall quality of life [26]. Therefore, the purpose of this review is to gather evidence on the current state of knowledge of potential long symptoms and consequences of COVID-19 in the pediatric population including physical, mental, behavioral, and social health, academic, and quality of life outcomes.

## METHODS

PRISMA guidelines were used as the search framework. A comprehensive search was completed via PubMed, Web of Science, Science Direct, medRxiv, and COCHRANE with the following search terms: Multisystem Inflammatory Syndrome or MIS-C, Pediatric Inflammatory Multisystem Syndrome Temporally Associated with SARS-Co-V-2 or PIMS-TS, COVID-19 and/or SARS-CoV-2, and children, adolescent, adolescence, or pediatric. PubMed filters applied: Abstract, Clinical Study, Clinical Trial, Comparative Study, Controlled Clinical Trial, Journal Article, Meta-Analysis, Observational Study, Randomized Controlled Trial, Review, Systematic Review, Humans, Child: birth-18 years, Newborn: birth-1 month, Infant: birth-23 months, Infant: 1-23 months, Preschool Child: 2-5 years, Child: 6-12 years, Adolescent: 13-18 years, from 11/1/2019 -/9/1/2021.

Studies were stratified by study setting and patient population type. Group A consisted of articles specifically mentioning MIS-C and/or PIMS-TS in their title, and/or their primary population diagnosed as MIS-C and/or PIMS-TS patients. Group B was defined by studies that took place within a hospital and/or participants having been studied for COVID-19 infection in any hospital department such as the ED, NICU, PICU, or ICU. If a population also included outpatients or was any study conducted outside of a hospital setting, then they were placed in Group C (Non-hospital). After removing duplicates and reading title and abstract, the number of articles were reported by articles found, those selected for literature review, and those specifically pertaining to or considered as relating to long-term effects, for primary review.

## RESULTS

The total number of articles selected for review was 130, with 34 deemed relevant or directly pertaining to long-term effects following a COVID-19 infection and/or effects of the COVID-19 pandemic in the pediatric population. In general, inclusion/exclusion characteristics were not well-defined, but there were five recent studies containing “long” or “long term” in their title.

### MIS-C, PIMS-TS and COVID-19 Acute Symptoms

Studies pertaining to MISC-C, PIMS-TS and/or acute COVID-19 symptoms are included in *Table 1*. While all of the common symptoms of COVID-19 such as fever, cough, shortness of breath, chills, muscle pain, headache, sore throat, and loss of taste or smell were reported in the pediatric studies reviewed, the symptoms seen consistently were fever and cough. There were additional respiratory symptoms reported across all pediatric population groups, such as sputum production, along with gastrointestinal (e.g., diarrhea), cardiovascular (e.g., cyanosis), and neurological (e.g., apnea) symptoms. One study examining hospitalized pediatric patients found that a portion demonstrated organ system failure [28]. Non-hospitalized patients in one study reported pain in the pharynx [38]. One study found that a portion of MIS-C pediatric patients reported/displayed (coryza) inflammation of the mucous membrane of the nasal cavity [34].

**Table 1.** Studies Reporting Acute Symptoms, MIS-C and PIMS-TS.

| **First Author and date of study** | **Age Group** | **Setting** | **Study Design/Time Frame** | **Acute Symptoms** | **Symptoms (Long, if any)** |
| --- | --- | --- | --- | --- | --- |
| Han et al., January 1<sup>st</sup>, 2021[27] | Patients younger than 19 years | Hospitals and Isolation Facilities across Korea | Case series (Feb 18<sup>th</sup> to March 31<sup>st</sup>, 2020) | Fever, respiratory symptoms, gastrointestinal symptoms, abdominal pain, and loss of smell or taste | Not reported |
| Zachariah et al., October 1<sup>st</sup>, 2020 [28] | Patients Less than or equal to 21 years | Tertiary care academically affiliated children's hospital in New York City | Retrospective (March 1<sup>st</sup> to April 15<sup>th</sup>, 2020) | Fever, respiratory, or gastrointestinal symptoms | Not reported |
| Verma et al., October 1<sup>st</sup>, 2020 [29] | Newborns | 4 major Metropolitan area hospitals, NYC | Observational (March 1<sup>st</sup> to May 10<sup>th</sup>, 2020) | One patient tested positive | Not reported |
| Shekerdemian et al., May 11<sup>th</sup>, 2020 [30] | Children between ages of 4 and 16 | PICUs across North America | Cross-sectional (March 14<sup>th</sup> and April 3<sup>rd</sup>, 2020) | Respiratory symptoms | Not reported |
| Zeng et al., July 1<sup>st</sup>, 2020 [8] | Neonates | Wuhan Children's Hospital | Cohort (Jan to Feb, 2020) | Shortness of breath | Not reported |
| DeBiasi et al., May 13<sup>th</sup>, 2020 [31] | Children and young adults | Children's National Hospital, Washington, DC | Observational retrospective cohort (March 15 and April 30, 2020) | respiratory symptoms (rhinorrhea, congestion, sore throat, cough, or shortness of breath), fever, headache, diarrhea, vomiting, loss of sense of smell or taste | Not reported |
| Leibowitz et al., | Febrile | Cohen Children's | Retrospective | Those infected were more | Not reported |
| February 1<sup>st</sup>, 2021 [32] | Infants less than 57 days of age | Medical Center of Northwell Health | (March 1<sup>st</sup> to April 30<sup>th</sup> of 2018, 2019, and 2020) | likely to display lethargy, feeding difficulties, and lower cell count |  |
| Mark et al., January 1<sup>st</sup>, 2021[33] | Infants younger than 3 months | N/A - Systematic review | Review (Nov. 1<sup>st</sup>, 2019 to June 15<sup>th</sup>, 2020) | Fever, respiratory, gastrointestinal, cardiac, and neurologic findings | Not reported |
| Xiong et al., September 1<sup>st</sup>, 2020 [34] | Less than or equal to 18 years | Princess Margaret Hospital in Hong Kong and Wuhan Children's Hospital in Wuhan | Cross-sectional (March 1<sup>st</sup> to April 30<sup>th</sup>, 2003 & Jan 21<sup>st</sup> to March 20<sup>th</sup>, 2020) | Fever, chills, myalgia, malaise, coryza, sore throat, sputum production, nausea, headache and dizziness | Not reported |
| Chao et al., August 1<sup>st</sup>, 2020 [35] | Patients aged 1mo to 21 years | Children's Hospital at Montefiore, United States | Retrospective cohort (March 15<sup>th</sup> and April 13<sup>th</sup>, 2020) | Fever, cough, shortness of breath, severe sepsis and septic shock syndromes, and acute respiratory distress syndrome | Not reported |
| Parri et al., December 1st 2020 [20] | Children with a median age of 45 months; interquartile range of 4 months–10.7 years | Pediatric Emergency Departments in Italy | Retrospective cohort (March 3<sup>rd</sup>, to May 2<sup>nd</sup>, 2020) | Fever, dry or productive cough, rhinorrhea, apnea, cyanosis, headache, dehydration | Not reported |
| Kainth et al., October 1<sup>st</sup>, 2020 [36] | Patients aged <22 years | Steven and Alexandra Cohen Children's Medical Center at Northwell Health in NYC, United States | Retrospective cohort (January 23<sup>rd</sup> to April 23<sup>rd</sup>, 2020) | Fever, lower respiratory symptoms, and gastrointestinal symptoms | Not reported |
| Laws et al., January 1<sup>st</sup>, 2020 [37] | Adults, and Children aged 3-17 years | Utah and Wisconsin households in the United States | Prospective cohort study (March to May, 2020) | Headache, sore throat, fever, and loss of taste or smell | Not reported |
| Wu et al., July 1<sup>st</sup>, 2020 [38] | Children <18 years | Qingdao Women and Children's Hospital and Wuhan Children's Hospital China | Retrospective cohort (January 20<sup>th</sup> to February 27<sup>th</sup>, 2020) | Fever, cough, fatigue, chest congestion, anorexia, diarrhea, dyspnea, headache, expectoration, myalgia, pharyngalgia, and dizziness | Prolonged fecal shedding of SARS-CoV-2 RNA |
| Dong et al., June 1<sup>st</sup>, 2020 [4] | Children <18 years | Data from the National Notifiable Infectious Disease Surveillance System at the Chinese CDC, analyzed at Shanghai Children's Medical Center | Nationwide case series (January 16<sup>th</sup> to February 8<sup>th</sup>, 2020) | Fever, cough (dry or productive), wheezing, congestion of the pharynx, fatigue, myalgia, sore throat, runny nose, sneezing, nausea, vomiting, abdominal pain, diarrhea, acute respiratory distress syndrome or respiratory failure, shock, encephalopathy, myocardial injury or heart failure, coagulation dysfunction, and acute kidney injury | Not reported |
| Heald-Sargent et al., September 1<sup>st</sup>, 2020 [39] | Patients aged 1mo to 65 years | Inpatient, outpatient, emergency department, and drive-through testing sites at a pediatric tertiary medical center, | Cohort study (March 23<sup>rd</sup> to April 27<sup>th</sup>, 2020) | Increased viral nucleic acid in their upper respiratory tract compared with older children and adults | Not reported |
|  |  | Chicago Illinois,<br>United States |  |  |  |
| Dionne et al.<br>November 1<sup>st</sup>, 2020<br>[40] | Patients<br>aged 0-22<br>years | Boston Children's<br>Hospital | Retrospective<br>cohort (March 1<sup>st</sup><br>to May 30th,<br>2020) |  | ECG anomalies |
| Yonker et al.<br>December 1<sup>st</sup>, 2020<br>[41] | Patients<br>aged less<br>than 21<br>years | Massachusetts<br>General Hospital | Cross-sectional<br>(not stated) |  | Fever |

Specifically, MIS-C and PIMS-TS patients presented with Kawasaki disease-like features, shock, fatigue, fever and inflammation [11,12][42,43][44-45]. PIMS-TS studies consistently reported Myocarditis [13][46-48], with additional PIMS-TS and MIS-C studies reporting cardiovascular findings such as atrioventricular block, coronary artery abnormalities, raised cardiac inflammatory markers, and transient valve regurgitation [11][40][49]. One PIMS-TS study from India, suggested that all children are at risk of COVID-19 infection, even if they present with mild symptoms or no symptoms [45].

### Long-Term Symptoms and Effects on Physical Health

*Table 2* summarizes current studies in the literature focused on the long physical symptoms of COVID-19 illness in the pediatric population. To date, a few studies with limited sample sizes, focusing on the non-hospitalized pediatric population, found that frequently reported physical symptoms were reported on average three to six months after infection including fatigue, muscle and joint pain, headache, insomnia, respiratory problems, palpitations, difficulty in concentration, and sensory problems [17, 70-73]. Weight gain due to lack of exercise and atopic dermatitis triggered potentially by a lack of exposure to sunlight and the outdoors, have also been reported [51].

**Table 2.** Studies Reporting Long Term Physical Symptoms.

| First Author and date of study | Age Group | Setting | Time Frame | Long Symptoms |
| --- | --- | --- | --- | --- |
| Bahar et al., December 1<sup>st</sup>, 2020 [50] | Patients less than 22 years | Children's National Hospital, DC | Retrospective (March 13<sup>th</sup> to June 21<sup>st</sup>, 2020) | Long-term viral shedding |
| Wu et al., July 1<sup>st</sup>, 2020 [38] | Newborns to 15 years | Qingdao Women and Children's Hospital and Wuhan Children's Hospital | Retrospective (January 20<sup>th</sup> to February 27<sup>th</sup>, 2020) | Long-term viral shedding |
| Li et al. May 19<sup>th</sup>, 2020 [51] | Children <14 years | Children's Hospital, Zhejiang University School of Medicine in Zhejiang Province. | Retrospective (January 1, 2020 to March 31, 2020) | Weight gain and atopic dermatitis. |
| Noh et al., January 21<sup>st</sup>, 2020 [52] | Children and adolescents ≤19 years | Households in Northern Virginia, U.S. | Cross-sectional, observational study (July to October, 2020) | None specifically.<br>SARS-CoV-2 antibody rate was double the adult rate.<br>Robust immune response to the nucleocapsid antigen |
| Buonsenso et al., January 26<sup>th</sup>, 2021 [17] | Children ≤18-year-old | Fondazione Policlinico Univeersitario A. Gemelli IRCCS (Rome, Italy) | Cross-sectional (September 1<sup>st</sup>, 2020 to January 1<sup>st</sup>, 2021) | Long-term health symptoms 120 days after COVID-19 infection, including fatigue, muscle and joint pain, headache, insomnia, respiratory problems and palpitations |
| Buonsenso et al., January 20<sup>th</sup>, 2021 [53] | Adults, and Children younger than 18 years of age | Fondazione Policlinico Univeersitario A. Gemelli IRCCS (Rome, Italy) | Prospective cohort (May 25<sup>th</sup> to July 15<sup>th</sup>, 2020) | None specifically.<br>Similar rates of SARS-CoV-2 IgG in all age groups studied |
| Ludvigsson, March 2021 [70] | Children with median age of 12 | Sweden | Case report | All five children had fatigue, dyspnoea, heart palpitations or |
|  | years (range 9-15) |  |  | chest pain, and four had headaches, difficulties concentrating, muscle weakness, dizziness and sore throats. |
| Lopez et al., March 28<sup>th</sup>, 2021 [71] | Children with median age of 142 months (IQR 117.8–166.8) | Spain | Prospective cohort (March to June 2020) | Persistent low □ grade fever, intense asthenia and severe headache |
| Radtke et al., July 15<sup>th</sup>, 2021 [73] | Children with median age of 11 years (IQR 9-13) | 55 randomly selected schools in the canton of Zurich in Switzerland | Prospective cohort (October and November 2020 to March and April 2021) | The most frequently reported symptoms lasting more than 12 weeks among seropositive children were tiredness (3/109 [3%]), difficulty concentrating (2/109 [2%]), and increased need for sleep (2/109 [2%]). |
| Osmanov et al. July, 2021 [74] | Children $\leq 18$ years old | Z.A. Bashlyaeva Children's Municipal Clinical Hospital (Moscow, Russia) | Prospective cohort study (April 2, 2020 to August 26, 2020) | Persistent symptoms among which fatigue (53, 10.7%), sleep disturbance (36, 6.9%,) and sensory problems (29, 5.6%) were the most common. |

From the many studies examining COVID-19 acute symptoms, a few studies also reported on immunological findings such as long-term viral shedding, or longer duration of viral particle expulsion through daily activities such as talking, exhaling, and eating. Two retrospective studies, one from the U.S. and one from China, both reported long-term viral shedding [38][50]. There are also many studies regarding IgG levels and immunological responses following COVID-19 infection. For example, one cross-sectional study found that the SARS-CoV-2 IgG rate was double in the children population compared to the adult population [52]. Conversely, another study found similar antibody rates across all age groups [53].

### Long-Term Effects on Mental Health

Studies pertaining to the long-term effects on mental health are included in *Table 3*. The most common mental health issues reported in the pediatric population throughout the COVID-19 pandemic were anxiety and depression, and these were only reported in papers looking at the COVID-19 pediatric patient groups [54,55]. From two non-hospitalized cross-sectional studies in China, one examined mental health effects in primary schools and the other in junior and senior high schools, with both studies reporting anxiety and depression during home confinement during the first few months of 2020 [54,55]. Conversely, a study in the U.S. examined the experiences of children within households identifying as Chinese-American. The authors found poorer mental health statuses as associated with higher levels of perceived racial discrimination [24].

**Table 3:** Studies Reporting Long Term Mental Health Outcomes.

| First Author and date of study | Age Group | Setting | Time Frame | Symptoms (Long), if any |
| --- | --- | --- | --- | --- |
| Li et al., January 19<sup>th</sup>, 2021 [54] | Adolescents | Junior and Senior High Schools in Wuhan | Cross-sectional (March 30<sup>th</sup> to April 7<sup>th</sup>, 2020) | Anxiety and depression |
| Xie et al., September 1<sup>st</sup>, 2020 [55] | Students grades 2-6 | 2 primary schools in the Hubei province | Cross-sectional (February 28<sup>th</sup> to March 5<sup>th</sup>, 2020) | Anxiety and depression |
| Cheah et al., November 1<sup>st</sup>, 2020 [24] | Parents, and children aged 10-18 years | Households in the U.S. that self-identified as Chinese | Retrospective Cohort (March 14<sup>th</sup> to May 31<sup>st</sup>, 2020) | Higher levels of perceived racial discrimination were associated with poorer mental health |
| Gassman-Pines et al., October 1<sup>st</sup>, 2020 [21] | Parents of a child or children aged 2-7 years | Large U.S. city | Prospective Cohort (February 20<sup>th</sup> to April 27<sup>th</sup>, 2020) | Increase in parental reporting of daily negative moods |
| Lujiten et al., November 4<sup>th</sup>, 2020 [26] | Children and adolescents aged 8-18 years | Two Dutch representative samples of children and adolescents in The Netherlands | Cross-sectional.<br>Before COVID-19 (Dec2017-July2018) and during the COVID-19 lockdown (April/May 2020). | Significantly worse PROMIS T-scores on all domains<br>Depressive symptoms, severe anxiety, and mental and health complaints |
| Alves et al., October 23<sup>rd</sup>, 2020 [18] | Children aged 9-15 years | Virtual visits during "stay-at-home" measures in the United States | April 22<sup>nd</sup> to July 29<sup>th</sup>, 2020 | Anxiety scores more than 5 standard deviations greater than values from healthy pediatric populations prior to the pandemic |

The daily moods of children were more frequently reported as negative during the pandemic as compared to before [21]. However, children that engaged in more physical activity during the pandemic reported less states of anxiety in a preliminary 2020 study in the U.S. [18]. A non-hospitalized, cross-sectional study in The Netherlands found significantly worse PROMIS T-scores on all domains, when comparing data from 2017-18 to data collected during April and May 2020 [26]. Mental health effects associated with the COVID-19 pandemic included depressive symptoms, severe anxiety, and patient-specific mental and social health complaints [26].

### Long-Term Effects on Social and Behavioral Health

Studies pertaining to the long-term effects on social and behavioral health are included in *Table 4*. For hospitalized neonates, the only long-term effects on behavioral health reported by parents were feeding, such as difficulties with or refusal to feed [20]. For older children, behavioral symptoms reported included clinginess, distraction, irritability, and fear of asking questions about the epidemic [19]. Other findings related to mood and emotional status included increases in being affectionate, restless, and frustrated [25]. The behavioral health of non-hospitalized children with COVID-19 had been reported by parents to have been worsening as the pandemic was progressing [56].

**Table 4.**
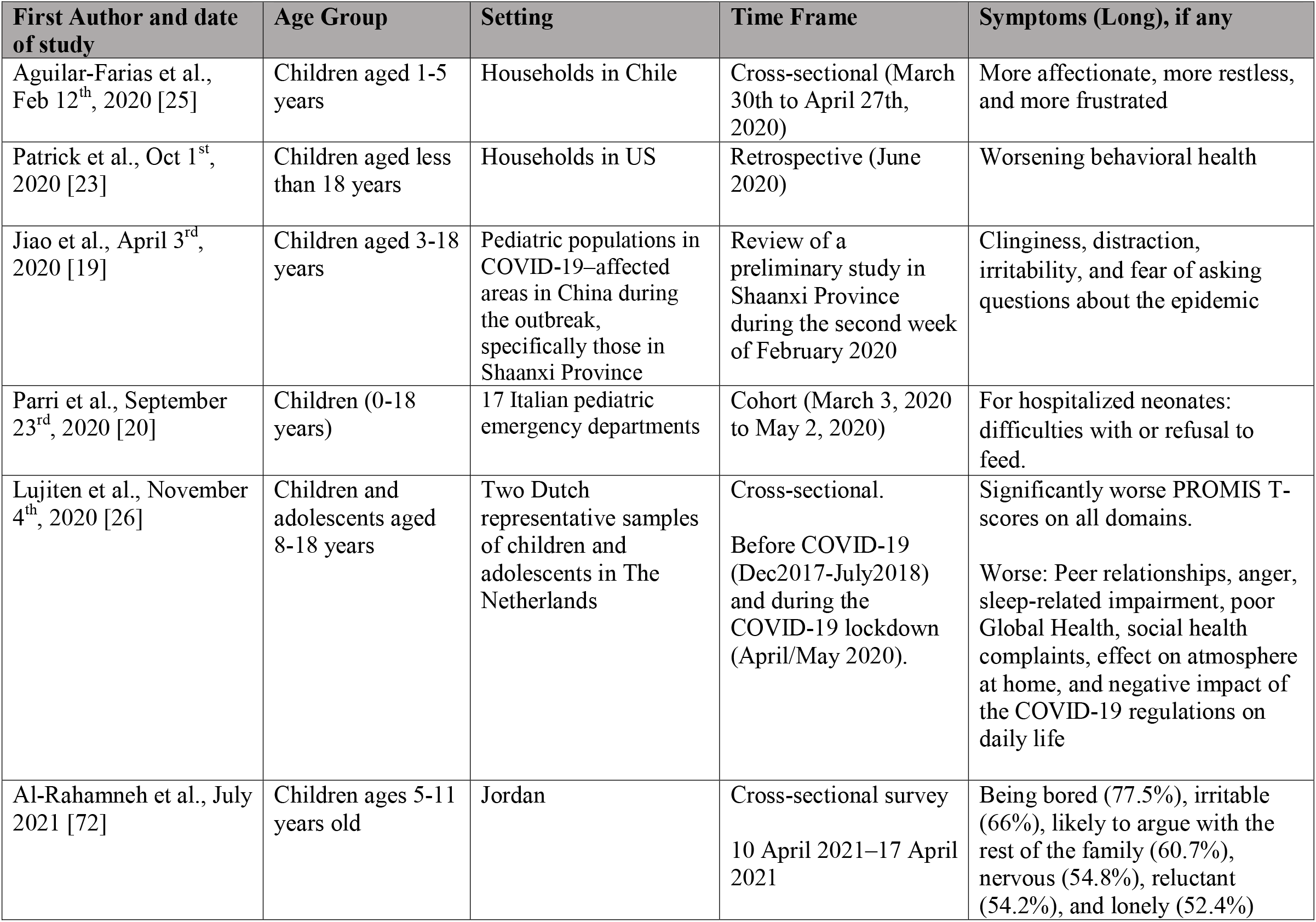
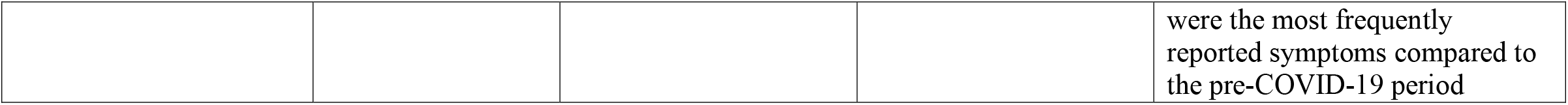
Studies Reporting Long Term Social/Behavioral Symptoms.

Additionally, a study among adolescents aged 8-18 years in The Netherlands has reported significant worse PROMIS T-scores on all domains including peer relationships, anger, sleep-related impairment, poor Global Health, social health complaints, effect on atmosphere at home, and negative impacts of the COVID-19 regulations on daily life [26].

### Long-Term Effects on Academics/Child Care

Long-term effects on academic performance and learning outcomes were not found in this review. The majority of articles regarding effects on school/childcare in general, focused primarily on changes in the organizational environment (i.e., in-person, virtual) and differences in COVID-19 infection rates. The locality, setting, duration, and stage of the pandemic that the study was conducted, were all limiting factors when comparing and contrasting the studies. For example, for childcare programs, it was suggested that findings should only be interpreted within the context of transmission rates and the infection mitigation efforts implemented by each program [57]. Throughout 2020, parents reported loss of child care alongside worsening parental mental health and child behavioral health [23]. In schools, there was an increase in the lack of access to technology and internet [22]. There was also an important precedence for developing school reopening plans to protect students who are most vulnerable to learning loss or reduced access to basic needs [22].

Other studies found varying associations in transmission rates among school setting/delivery while some took into account trends before and after school re-openings. A Florida county-level study found a 1.2-fold increase in COVID-19 infection rates among elementary schools, 1.3 in high schools and no effect for virtual learning, after school re-openings [58]. Conversely, a national study in Italy one month after school re-openings found low transmission in schools, mainly among younger students [59]. A U.S. state-level study found an increased prevalence of COVID-19 in adolescents and youth compared to adults in the summer of 2020 [23].

### Long-Term Effects on Quality of Life and Social Determinants of Health

Studies pertaining to long-term effects on Quality-Of-Life (QOL) and the Social Determinants of Health (SDOH) were not outlined in a table, as these outcomes relate more to parents and changes in environmental settings. Many articles focused on examining quality of life issues such as nutrition, home environment, overall well-being, daily moods, and mental/emotional attitudes toward the pandemic and quarantine measures. They also analyzed effects on social determinants of health such as insurance status, healthcare needs, food insecurity, housing, income status, and caregiving burdens. These outcomes were mostly measured from parental surveys and questionnaires completed by parents or caregivers. In a non-hospital study, authors found an increase in food insecurity, nutrition barriers, homelessness or use of temporary housing [22]. Similarly, another study found an increase in moderate to severe food insecurity, alongside changes in insurance status [23]. One parent survey found an increase in frequency of parent-reported daily negative mood [21]. This article also found that the parents’ and children’s well-being was strongly associated with the number of reported hardships [21]. Hardships included job loss, income loss, caregiving burden, illnesses, etc.

Preliminary reports found similar findings in worsening quality-of-life issues and increased health inequities among social determinants of health. Parents experienced anxiety, changes and limitations to healthcare access, and overall “collateral” damage to their well-being as a result of economic impacts and social isolation [60]. A cross-sectional study on children and adolescents in the Netherlands found an increase in mental and social health complaints during the lockdown with the majority reporting a negative impact of COVID-19 on their life [26]. Families were concerned about the COVID-19 pandemic and quarantine measures, especially towards negative impacts on the economy [61]. These findings also raise concern regarding stability and safety within the home environment. The pandemic’s impact on child abuse and claims remains hidden, underscoring the need for further research in this field [62].

### COVID-19 and Influenza Vaccine Opinions

Three hospital studies examined parental opinions and interests regarding COVID-19 and influenza vaccines for their children. One study discussed the overall exacerbation of polarity in COVID-19 vaccination uptake, and those parents who originally chose to not vaccinate their children for influenza, were more likely to commit to this decision than parents who originally chose to vaccinate their children and vice versa [63]. Similarly, one preliminary study found that 60% of parent participants reported that they were likely to vaccinate their children, as well as themselves against COVID-19 [64]. Separately, another study found an increase in willingness to vaccinate against influenza [65].

## DISCUSSION

To the authors’ knowledge, this is one of the first scoping reviews focused on the long physical, psychological, behavioral, academic, and social consequences of COVID-19 disease and the pandemic in general in the pediatric population. From November 2019 to September 2021, our search found that out of approximately 130 publications, roughly 34 contained relevant information, and 5 specifically examined “long-hauler symptoms’’ or long-term effects in the pediatric population. One of the largest long COVID-19 study was a cross-sectional study from Italy examining long COVID-19 in a small sample (N=129) of children (≤18 years), with more than half of their patients reporting at least one long-term symptom [17]. Our search findings were also consistent with the systematic review by Ludvigsson et al., published March 2021 in Sweden. The authors reviewed 179 publications, deemed 19 relevant, and did not find any containing information on long COVID-19 in children [66]. They also included findings from their case report on five patients, who all presented with the primary persisting symptom of fatigue 6-8 months following a clinical COVID-19 diagnosis [66].

As of March 2021 and within the scope of our review search parameters, one study has now been published reporting physical long symptoms in children. These symptoms which commonly persist following a normal infection recovery include standard symptoms such as fever, cough, shortness of breath, muscle pain, and a headache. Some additional physical long symptoms observed in children were insomnia and heart palpitations. The persistence of these symptoms could possibly be attributed to SARS-CoV-2 triggering an abnormal immunological or inflammatory response in specific areas of the body that express the ACE2 receptor [38][50][67]. The invasion and persistence of SARS-CoV-2 in the central nervous system could also potentially be associated with the occurrence of mental health issues such as anxiety and depression, with further research needed to understand the mechanisms of action [67-69]. However, many long symptoms and their etiologies may be subjective in nature and therefore, additional research is needed to investigate the pathogenesis of long COVID-19 symptoms across all pediatric groups, including those clinically diagnosed with MIS-C and PIMS-TS.

The majority of reviewed studies pertaining to the long-term effects of COVID-19 in children focused on mental health, social/behavioral health and environmental outcomes as a result of quarantine and social distancing. The social impact of COVID-19, primarily the mandated stay-at-home orders in 2020 and continued social distancing protocols into 2021, continues to contribute a larger role in the maintenance of social/behavioral health and mental health disorders, or at minimum, their individual symptoms. Social interaction including familial and peer relationships is integral to the development, growth and learning environment of children. The physical and emotional interactions with other individuals, both of their own age and older, facilitates proper neural development, especially regarding impulse control, mood regulation and academic development. With the potential for physical symptoms to exacerbate psychological symptoms and all child age groups awaiting COVID-19 vaccine approval, further research is needed to determine the full course of SARS-CoV-2 in the pediatric body and any persisting long-hauler effects that could compromise quality of life, even if mild in severity.

## CONCLUSION

In contrast to earlier reports suggesting that the outcomes or physical effects of COVID-19 in the pediatric population were milder or less severe in comparison to the adult population, the findings from this review indicate that a subgroup of children are still at risk to develop more severe and long-term presentation of symptoms, even more so for children diagnosed with MIS-C, PIMS-TS and multiple organ system failure. In addition, COVID-19 has had a moderate impact on children and adolescents’ social environment, which may exacerbate current and future physiological, psychological, behavioral, and academic outcomes. The relative lack of evidence evaluating the long-term effects of the recent COVID-19 pandemic and infections on the pediatric population, suggests the need for more prospective studies examining the long-hauler effects of an initial infection, as compared to retrospective/cross-sectional studies examining symptom presentation. This review serves as a continuum in which further research is needed to thoroughly investigate and understand the complete effects, from acute to long-term, that SARS-CoV-2 induces in the human body, especially for the pediatric population.

## Data Availability

N/A

